# Late surges in COVID-19 cases and varying transmission potential partially due to public health policy changes in 5 Western states, March 10, 2020-January 10, 2021

**DOI:** 10.1101/2021.07.04.21259992

**Authors:** Xinyi Hua, Aubrey R. D. Kehoe, Joana Tome, Mina Motaghi, Sylvia K. Ofori, Po-Ying Lai, Sheikh Taslim Ali, Gerardo Chowell, Anne C. Spaulding, Isaac Chun-Hai Fung

**Author notes:** Corresponding author: Isaac Chun-Hai Fung, PhD. Xinyi Hua, Aubrey R. D. Kehoe, Joana Tome and Mina Motaghi serve as co-first authors.

## Abstract

**Objective:** This study investigates how the SARS-CoV-2 transmission potential varied in North Dakota, South Dakota, Montana, Wyoming, and Idaho from March 2020 through January 2021.

**Methods:** Time-varying reproduction numbers, *R*_*t*_, of a 7-day-sliding-window and of non-overlapping-windows between policy changes were estimated utilizing the instantaneous reproduction number method. Linear regression was performed to evaluate if per-capita cumulative case-count varied across counties with different population size.

**Results:** The median 7-day-sliding-window *R*_*t*_ estimates across the studied region varied between 1 and 1.25 during September through November 2020. Between November 13 and 18, *R*_*t*_ was reduced by 14.71% (95% credible interval, CrI, [14.41%, 14.99%]) in North Dakota following a mask mandate; Idaho saw a 1.93% (95% CrI [1.87%, 1.99%]) reduction and Montana saw a 9.63% (95% CrI [9.26%, 9.98%]) reduction following the tightening of restrictions. High-population counties had higher per-capita cumulative case-count in North Dakota at four time points (June 30, August 31, October 31, and December 31, 2020). In Idaho, North Dakota, and South Dakota, there was a positive correlation between population size and per-capita weekly incident case-count, adjusted for calendar time and social vulnerability index variables.

**Conclusions:** *R*_*t*_ decreased after mask mandate during the region’s case-count spike suggested reduction in SARS-CoV-2 transmission.

## INTRODUCTION

A case of coronavirus disease 2019 (COVID-19) reported in the state of Washington on January 21, 2020 heralded the arrival of the pandemic in the United States (U.S.), but neighboring states were spared for over a month. While Idaho borders Washington, it and four other contiguous western states—North Dakota, South Dakota, Montana, and Wyoming—did not report a single case until 49 days later.^1^ All five states are sparsely populated.^2^ Due to demographic heterogeneity, and the division of power between federal and state governments, the reaction to various phases of the pandemic have differed in type and timing by region, resulting in dissimilar patterns of disease spread around the country.^3^ More regional studies are needed to assess the variation in the spatial heterogeneities of the transmission of the severe acute respiratory syndrome coronavirus 2 (SARS-CoV-2) and the distribution of disease burden. In this paper, we will analyze the disease pattern in North Dakota, South Dakota, Idaho, Montana and Wyoming to provide insight for epidemiologists and to help policy makers with future decision-making processes.

Orders from the executive branch of federal and state governments and state health officer orders regarding the COVID-19 pandemic included social distancing, quarantine and isolation, mask mandates, closure of businesses, and COVID-19 testing requirements. These were implemented in different time frames across this 5-state region. Evaluating the existing data to explore the impact of policy implementation on COVID-19 pandemic may provide insight on the policies with high impact in reducing the transmission of SARS-CoV-2 infection, morbidity and mortality. Quantifying epidemiologic characteristics of the COVID-19 pandemic in these states, so that we can document the potential effect of policies and non-pharmaceutical interventions that reduce COVID-19 transmission and mortality, may make us better prepared for the emergence of future infectious disease epidemics.

Central to the description of an epidemic’s transmission potential is the reproduction number. The basic reproduction number, also called *R*_*0*_, shows the transmissibility of an infectious agent at the beginning of an outbreak; it is calculated as the average number of secondary cases generated by a primary case in a completely susceptible population, prior to any behavioral changes or public health interventions.^4^ On the other hand, the time-varying reproduction number, also known as *R*_*t*_, is a time-dependent estimate of the average number of secondary cases that are generated from one case at time *t*, after there are behavioral changes, depletion of the susceptible population, and implementation of disease control policies.^5,6^

An *R*_*t*_ larger than one indicates sustained transmission and the epidemic is expected to expand in the population. An *R*_*t*_ less than one indicates that the epidemic will tend to decline. Therefore, it is used as an indication of the effectiveness for infection control measures.^4,5^ Calculating *R*_*t*_ over the course of pandemic, from March 2020 through January 2021, this study aims to investigate the time-dependent variability in transmission potential of SARS-CoV-2 in these five states in different time periods, and explore their relationship with the changes in the states’ public health policies.

## METHODS

We used time-series data for the COVID-19 pandemic during March 10, 2020 – January 10, 2021, in the states of North Dakota, South Dakota, Idaho, Montana, and Wyoming. A detailed list and description of all counties (North Dakota’s 53 counties, South Dakota’s 66 counties, Idaho’s 44 counties, Montana’s 56 counties, and Wyoming’s 23 counties) are provided in **Supplementary Table 1**.

### DATA ACQUISITION

We downloaded the cumulative confirmed case count during March 10, 2020 – January 10, 2021, for all five states, including the counties located in each state from the New York Times GitHub data repository.^7^ The first case of each state was reported during the same week, on March 10 (South Dakota), March 11 (North Dakota and Wyoming), and March 13 (Idaho and Montana) respectively. Our cutoff point for all five states was January 10, 2021. Our timeframe covered nearly ten months from the first reported case in those states. We obtained the daily number of newly confirmed COVID-19 cases from the reported cumulative case count numbers (**Appendix A**). We also retrieved 2019 county-level estimated population data for all five states from the U.S. Census Bureau.^8^

We collected and assessed the executive orders from the governors’ offices of the five states and identified the timing of the orders to implement and the announcements permitting relaxation of public health interventions in each state respectively (**Supplementary Table 2**).

### STATISTICAL ANALYSIS

We estimated the time-varying reproduction number, *R*_*t*_, using the instantaneous reproduction number method with parametric definition of the serial intervals as proposed by Cori et al.^6^ as implemented in the R package ‘EpiEstim’ version 2.2-3. The instantaneous reproduction number is one of a few definitions of *R*_*t*_. It is an estimate of the transmissibility of the disease at current time *t*, assuming that it is the same as the transmissibility of prior cases that result in the number of their secondary cases at current time *t*. An average *R*_*t*_ can be estimated using a fixed sliding window or non-overlapping time windows defined by the user. *R*_*t*_ is a time varying measure of transmissibility and defined as the ratio between the number of incident cases at the time *t*, and the total infectiousness of all infected individuals at the time *t* accounted during their infectious periods. As county-level data may give information of health inequities, or crucial information in differences in COVID-19 transmission rate,^9^ we conducted our analysis based on county level data. We first reconstructed the date of infection according to Gostic et al.,^10^ by shifting the time series by nine days backward (assuming a mean incubation period of 6 days and a median delay to testing of 3 days).^11^ We assumed the serial interval distribution with a mean of 4.60 days and a standard deviation of 5.55 days.^12^ Besides using the default 7-day sliding window, we also estimated *R*_*t*_ by the non-overlapping time periods when different combinations of non-pharmaceutical interventions (i.e., face masking, social distancing, school and business closure, etc.) have been implemented (we call them policy change *R*_*t*_ thereafter). The policy change *R*_*t*_ is the average of the daily *R*_*t*_ over the non-overlapping time period between two major policy changes.

Percentage change is often utilized to help identify the magnitude and direction of change in a statistic. We calculated percentage change for both the 7-day sliding window *R*_*t*_ and the policy change *R*_*t*_ (**Table 1**). This was calculated utilizing percentage change 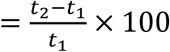, at the date of each policy implementation and face-to-face school resumption. For 7-day sliding window *R*_*t*_ each date of interest is considered time 2 (t_2_) and the previous 7-day period was utilized as time 1 (t_1_); for the policy change *R*_*t*_ each time window was compared to the previous window. We utilized EpiEstim “sample from the posterior R distribution” function to sample 1000 estimates of *R*_*t*_ for each t_1_ and t_2_ then estimate the 95% credible intervals (2.5 and 97.5 percentile) of the calculated percentage change through bootstrapping.

**Table 1:** Percentage change of R_t_ and 95% credible intervals (CrI) at policy implementation and important dates calculated with both 7-day sliding window and non-overlapping window.

| State | Date (2020) |  | Percentage Change and 95% CrI |  |  |  |
| --- | --- | --- | --- | --- | --- | --- |
|  |  |  | 7-Day Sliding Window |  | Non-Overlapping Window |  |
| North Dakota | 16-Mar | Policy A | 8.86% | [-22.86%, 52.67%] | -18.01% | [-39.68%, 10.12%] |
|  | 30-Mar | Policy B | -10.96% | [-15.3%, -10.46%] | -21.43% | [-32.18%, -8.84%] |
|  | 26-May | Policy C | 6.98% | [-1.61%, 16.12%] | 4.30% | [0.47%, 8.19%] |
|  | 7-Sep | School Open** | -8.03% | [-8.23%, -7.83%] |  |  |
|  | 21-Sep | Policy D | 0.17% | [0.01%, 0.33%] | 0.82% | [-0.82, 2.49%] |
|  | 13-Nov | Policy E | -14.71% | [-14.99%, -14.41%] | -27.27% | [-28.24%, -26.09%] |
|  | 21-Dec | Policy F | 32.44% | [26.53%, 38.09%] | 20.85% | [16.86%, 25.10%] |
| South Dakota | 13-Mar | Policy A | 94.16% | [-9.05%, 404.87%] | 89.99% | [7.91%, 285.91%] |
|  | 6-Apr | Policy B | -38.85% | [-43.80%, -33.50%] | -42.62% | [-47.18%, -38.20%] |
|  | 28-Apr | Policy C | 37.34% | [24.20%, 51.22%] | 9.64% | [7.00%, 12.28%] |
|  | 13-Aug | Policy D | 16.47% | [7.06%, 26.37%] | 7.59% | [7.09%, 8.08%] |
|  | 7-Sep | School Open** | 12.36% | [8.07%, 16.72%] |  |  |
|  | 25-Sep | Policy E | 18.84% | [18.19%, 19.48%] | -10.38% | [-12.00%, -8.86%] |
|  | Nov 13-18 | Average* | 2.80% | [-7.85%, 34.52%] |  |  |
| Idaho | 25-Mar | Policy A | -59.94% | [-60.57%, -59.31%] | -62.67% | [-65.75%, -59.49%] |
|  | 1-May | Policy B | 11.14% | [10.38%, 11.98%] | 41.97% | [35.64%, 48.05%] |
|  | 13-Jun | Policy C | 8.68% | [-0.84%, 19.05%] | -5.02% | [-9.42%, -0.83%] |
|  | 7-Sep | School Open** | 13.14% | [12.52%, 13.73%] |  |  |
|  | 27-Oct | Policy D | 2.60% | [2.27%, 2.91%] | -2.67% | [-4.09%, -1.24%] |
|  | 14-Nov | Policy E | -1.93% | [-1.99%, -1.87%] | -6.24% | [-7.64%, -4.83%] |
| Montana | 13-Mar | Policy A | -3.99% | [-38.99%, 49.24%] | -51.24% | [-66.21%, -27.83%] |
|  | 7-May | Policy B | -14.92% | [-65.10%, 107.88%] | 22.60% | [10.14%, 35.95%] |
|  | 15-Jul | Policy C | -12.43% | [-12.55%, -12.31%] | -16.17% | [-17.27%, -15.08%] |
|  | 1-Sep | Policy D | 34.83% | [33.82%, 35.85%] | 8.52% | [5.54%, 11.28%] |
|  | 7-Sep | School Open** | 20.73% | [10.90%, 30.98%] |  |  |
|  | 18-Nov | Policy E | -9.63% | [-9.98%, -9.26%] | -17.58% | [-18.75%, -16.38%] |
| Wyoming | 13-Mar | Policy A | 15.46% | [-29.23%, 91.29%] | -32.95% | [-54.24%, 2.84%] |
|  | 28-Apr | Policy B | -29.99% | [-46.75%, -6.23%] | -5.13% | [-5.79%, -4.48%] |
|  | 10-Jun | Policy C | 1.17% | [-1.03%, 3.64%] | 5.69% | [-4.57%, 16.99%] |
|  | 13-Jul | Policy D | 0.30% | [-0.76%, 1.43%] | 1.96% | [-2.99%, 6.41%] |
|  | 7-Sep | School Open** | 19.56% | [1.66%, 39.27%] |  |  |
|  | 14-Oct | Policy E | -1.98% | [-2.64%, -1.33%] | -8.84% | [-11.03%, -6.78%] |
|  | Nov 13-18 | Average* | -13.22% | [-29.76%, 21.92%] |  |  |
|  | 7-Dec | Policy F | 10.58% | [10.34%, 10.83%] | -13.25% | [-15.30%, -10.97%] |

\*Note: neither South Dakota nor Wyoming had any policy change during this time-period of interest so bootstrapping was performed on the average of the 6-day range of policy implementation from the other three states.
\*\*Note: As of September 7<sup>th</sup>, 2020, all states had resumed face-to-face K-12 education.

As a note, *R*_*t*_ percentage change was calculated for the fall implemented interventions of North Dakota on November 13 (Mask order), Idaho on November 14 (Stay Health Order-Stage 2: Gatherings of no more than 10 people, and vulnerable population were strongly recommended to stay at home), and Montana on November 18 (Additional mitigation measures were implemented including reduced size of gatherings to no more than 25). Since neither South Dakota nor Wyoming ever ordered interventions during the same time period, for comparison purposes we calculated the 7-day-sliding-windows *R*_*t*_ percentage change for both states with November 13-18, 2020 as t_2_ and November 6-11, 2020 as t_1_.

We assessed the power-law relationship between the cumulative case count of COVID-19 and the population size of the counties. Cumulative case count would be exactly proportional to population size if the per capita cumulative case count remained the same across all counties of different population size (**Appendix B**). We ran a linear regression model between log_10_-transformed per capita cumulative case count and log_10_-transformed population size. Counties with lower population sizes would have a higher per capita cumulative case count if the slope of the regression line was negative, and a lower per capita cumulative case count if the slope was positive.^13-16^ We conducted the log-linear regression considering four different dates: June 30^th^, August 31^st^, October 31^st^ and December 31^st^, 2020.

To further explore the relationship between the per capita weekly incident case count and county’s population size, log linear analysis was performed as univariable and multivariable analysis. Multivariable models were adjusted for time (more specifically a categorical variable of each 7-day period from March 2020 to January 2021) and other factors that may impact this association. Those other factors were variables that we chose from the CDC Social Vulnerability Index (SVI), which was created by Geospatial Research, Analysis & Services Program to identify and map the most vulnerable communities with a need for support in emergency events.^17^ CDC utilizes the U.S. census data to estimate the social vulnerability for every census tract and every county in each state. The SVI is composed of many variables which are categorized in four different themes (socio-economic status, household composition and disability, race/ethnicity/languages, and housing type/transportation). In times of COVID-19, it becomes meaningful to account for social vulnerability variables when assessing the relationship between weekly incident case count per capita and population size. Those variables may partially explain the variability of weekly incident case count per capita. Among many variables included in the SVI database, we chose those variables we thought were more representative and had less collinearity with each other. The chosen variables were the percentage of people living below the poverty threshold, the percentage of people without a high school diploma, the percentage of people 65 and older, the percentage of the civilian non-institutionalized population with a disability, the percentage of minorities (all people excluding white, non-Hispanic) estimate, and the percentage of occupied housing units with more people than rooms estimate. We obtained our data from the Agency for Toxic Substances and Disease Registry.^18^

All statistical analyses were performed using R version 4.0.3 (R Core Team, R Foundation for Statistical Computing, Vienna, Austria). Maps (**Figure 1 and Figure 2**) were created using R version 3.5.1 (R Core Team, R Foundation for Statistical Computing, Vienna, Austria). The significance level was set a priori at α=0.05.

**Figure 1.**
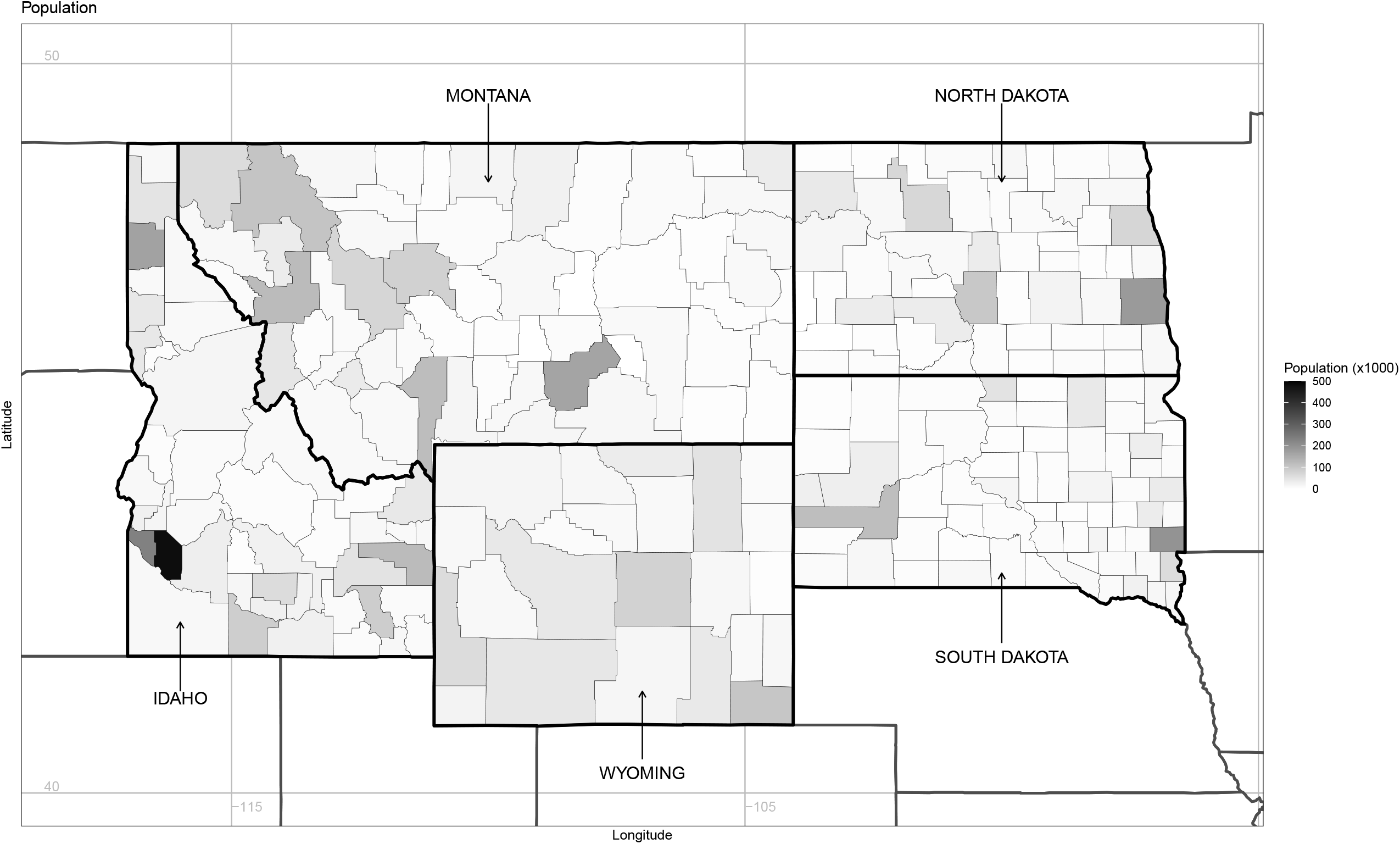
County-level population map (2019) of North Dakota, South Dakota, Idaho, Montana, and Wyoming, USA.

**Figure 2.**
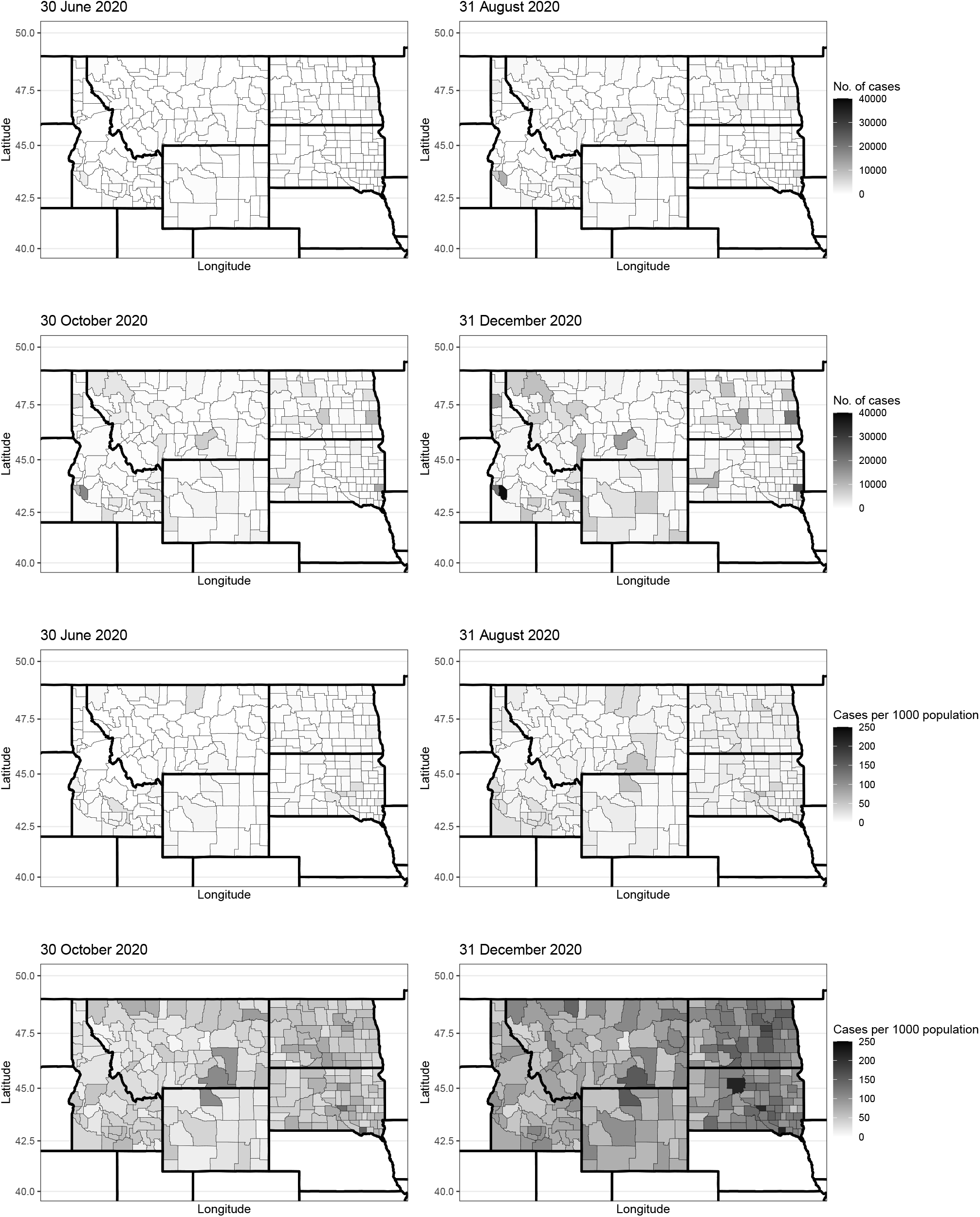
County-level maps of North Dakota, South Dakota, Idaho, Montana, and Wyoming by cumulative case count (top four maps), and cumulative case counts per 1000 population (bottom four maps) on June 30, August 31, October 31, and December 31, 2020 (date of report).

### ETHICS

The Georgia Southern University Institutional Review Board made a non-human subject determination for this project (H20364) under the G8 exemption category according to the Code of Federal Regulations Title 45 Part 46.

## RESULTS

### *R*_*t*_ ESTIMATES AT THE STATE LEVEL

From March 10, 2020 to January 10, 2021, the daily number of new cases showed at least one peak in the Fall across all five states, except Idaho which had a summer peak (**Figure 3, left panel**). All five states had a very similar qualitative trajectory. For instance, as of January 10, 2021, Idaho had reported 149,742 cases, the highest cumulative numbers of COVID-19 cases among five states. South Dakota had reported 113,318 cases, North Dakota 94,724 cases, Montana 86,324 cases, and Wyoming 46,832 cases. **Figure 2** presents the geospatial dynamics of cumulative cases number and cumulative case number per 1000 population (incidence) by county in five states at four different dates of report between March 10, 2020 and January 10, 2021: June 30^th^, August 31^st^, October 31^st^, and December 31^st^, respectively.

**Figure 3.**
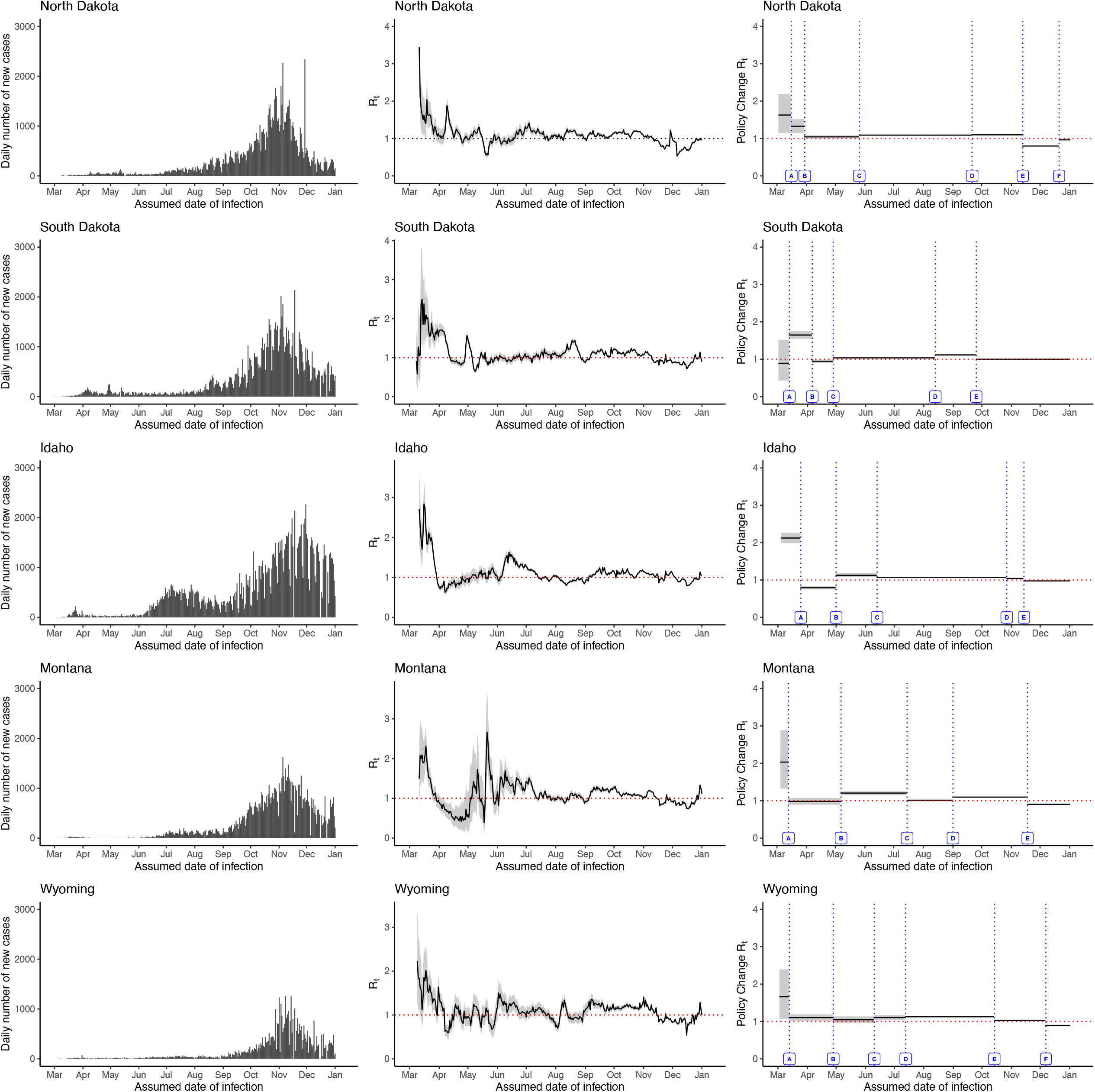
The daily number of new cases by their assumed date of infection (left panel), 1-week sliding window *R*_*t*_ estimates (middle panel), and non-overlapping window *R*_*t*_ estimates by policy change (right panel) of North Dakota, South Dakota, Idaho, Montana, and Wyoming, USA, March 10, 2020 – January 10, 2021 (date of report). **North Dakota**: A = Schools closed; B = Promote physical distancing; C = Testing order; D = Lifted travel and quarantine order; E = Mask order; F = Allow all restaurants and bars to resume normal hours. **South Dakota**: A = State of Emergency declared; B = Minnehaha and Lincoln stay at home order; C = All citizens shall implement ‘back to normal plan’; D = All K-12 schools were mandated to start in-person classes again; E = long-term facilities began to relax visitor restrictions. **Idaho**: A = Statewide stay home order; B = Stay Healthy Order-Stage 1. Businesses and governmental agencies resumed operations at physical locations; C = Stay Health Order-Stage 4. Gatherings of any size were allowed and non-essential travel could resume; D = Stay Health Order-Stage 3. Patrons of bars, nightclubs, and restaurants must remain seated. Face coverings were required at long-term care facilities; E = Stay Health Order-Stage 2. Gatherings of no more than 10 people, and vulnerable population were strongly recommended to stay at home. **Montana**: A = State of Emergency declared; B = School reopened; C = Statewide mask requirement; D = School’s Fall semester started; E = Additional mitigation measures were implemented including reduced size of gatherings to no more than 25. **Wyoming**: A = All businesses and schools were closed; B = Gymnasiums and childcare facilities were reopened; C = K-12 schools, colleges, universities, and trade schools resumed on-site instructions; D = Removed some conditions and restrictions were applicable to restaurants; E = Additional mitigation measures were implemented including reduced size of gatherings to no more than 50; F =Required face coverings in certain places with exceptions.

The 7-day-sliding-window *R*_*t*_ estimates were between 2 and 3, in the beginning of the pandemic across all five states. In Idaho, Montana, and Wyoming, the 7-day-sliding-window *R*_*t*_ estimates dropped below 1 in the early-April; and in North Dakota and South Dakota, the 7-day-sliding-window *R*_t_ estimates briefly drop below 1 in mid-May. Then the 7-day-sliding-window *R*_*t*_ estimates steadily stayed above 1 between September and December which correspond to the fall/winter surges in five states. At the end of the study period, the 7-day-sliding-window *R*_*t*_ estimates were slightly above 1 (Idaho, Montana, and Wyoming) or around 1 (North Dakota and South Dakota), demonstrating the extensive community transmission of SARS-CoV-2 in those states.

As of September 7, 2020 all students in K-12 grades resumed face-to-face instruction in all five states. Compared to the week prior, the 7-day-sliding-window *R*_*t*_ increased in Montana, Wyoming, Idaho, South Dakota; whereas North Dakota saw a reduction in *R*_*t*_ (**Table 1 & Figure 3, right panel**). Thus, these five states experienced an average of 11.55% increase in the 7-day-sliding-window *R*_*t*_.

Following the region’s fall surge in cases, North Dakota implemented a mask mandate on November 13^th^ (Policy E) which contributed to a 14.71% decrease in *R*_*t*_. Idaho saw a 1.93%, and Montana a 9.63% decrease in *R*_t_ following the implementation of stricter policies (Policy E respectively). Meanwhile in mid-November 2020, neither Wyoming nor South Dakota implemented any additional restrictions seeing differing effects on *R*_*t*_; Wyoming’s *R*_*t*_ decreased by 13.22% where South Dakota’s *R*_*t*_ increased by 2.8%.

Percentage change in policy change *R*_*t*_ were computed by comparing each time window (between each major policy change) to the previous (**Table 1**). Detailed description of *R*_*t*_ for each state can be found in **Appendices C and D**.

### POWER-LAW RELATIONSHIP BETWEEN CUMULATIVE CASE NUMBER AND POPULATION SIZE

**Figure 4** presents a linear regression relationship between the log_10_-transformed per capita cumulative case number and the log_10_-transformed population size for a total of 242 counties in North Dakota, South Dakota, Idaho, Montana, and Wyoming. Each panel corresponds to an assessed date (date of report), June 30^th^, August 31^st^, October 31^st^, and December 31^st^, 2020, respectively; each regression line represents a state in a specific assessed date. The slope, *m*, and its 95% confidence internal of every regression line are presented in **Supplemental Table 3**. North Dakota was the only state that had significant slopes at all four time points (*m*=0.2758, 0.2171, 0.0729, 0.0986; *p*=0.0034, 0.0018, 0.046, 0.0024; respectively). Overall, the slopes of regression lines at four different assessed dates were found slightly positive in June and August assessments but very close to zero in October and December assessments as the pandemic unfolded, except that of Montana on June 30^th^, 2020. This meant towards the end of 2020, there was no heterogeneity of per capita cumulative case count across 242 counties with different population size, suggesting an extensive community transmission of SARS-CoV-2 as the pandemic progressed. However, as we mentioned previously, the slope of Montana’s June regression line (−0.1423) was slightly less than zero. When *m*<0, it suggests counties with lower population sizes would experience a higher per capita cumulative case count. In other words, the rural counties in Montana experienced a slightly higher impact of the COVID-19 pandemic than other counties in the same state by mid-2020. **Figures S1-S5 (Appendix E)** present the same regression analyses, separately for each state, with outliers in log-transformed per capita cumulative case number highlighted.

**Figure 4.**
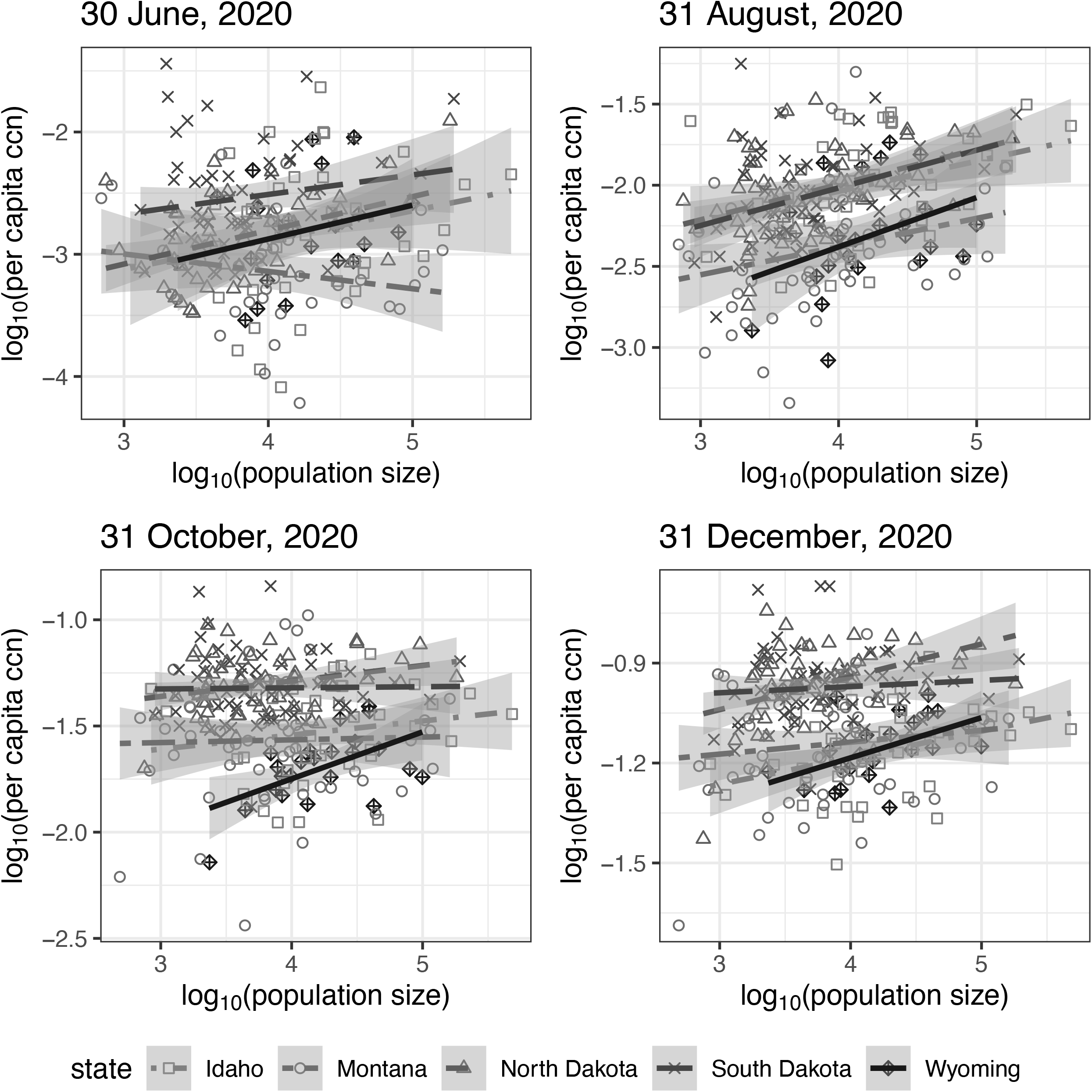
Linear regression between log_10_-transformed per capita cumulative case number (ccn) and log_10_-transformed population size by county (grouped in states) for North Dakota, South Dakota, Idaho, Montana, and Wyoming on June 30^th^, August 31^st^, October 31^st^, and December 31^st^, 2020 (date of report).

### MULTIPLE REGRESSION ANALYSIS BETWEEN PER CAPITA WEEKLY INCIDENT CASE COUNT AND POPULATION SIZE, ADJUSTED FOR SVI VARIABLES

Linear regression results for evaluating the relationship between log_10_-transformed per capita weekly incident case count and log_10_-transformed population size is shown in **Table 2**. Each ten-fold increase in population size, after adjusting for the time of data collection (therefore adjusting for different policies running in the state or different stages of the pandemic), was associated with an increase of 0.06—0.27 in the log_10_-transformed per capita weekly incident case count, in the studied states and all these changes were statistically significant. After controlling for SVI confounders, in addition to time, a positive trend was found in the association between population size and per capita weekly incident case count in Idaho, North Dakota, and South Dakota. However, these coefficients were not statistically significant for Montana and Wyoming.

**Table 2.** The linear regression analysis between log_10_-transformed per capita weekly incident case count and log_10_-transformed population size for North Dakota, South Dakota, Idaho, Montana, and Wyoming, March 2020 -January 2021.*

|  | Unadjusted Model |  | Adjusted for Weeks |  | Adjusted for all Variables† |  |
| --- | --- | --- | --- | --- | --- | --- |
|  | Parameter Estimate | (95% CI) | Parameter Estimate | (95% CI) | Parameter Estimate | (95% CI) |
| North Dakota | 0.066 | (-0.009,0.141) | 0.190 | (0.148, 0.233)* | 0.195 | (0.131, 0.259)* |
| South Dakota | -0.018 | (-0.092,0.056) | 0.143 | (0.099, 0.187)* | 0.127 | (0.069, 0.185)* |
| Idaho | 0.005 | (-0.071,0.080) | 0.155 | (0.108, 0.201)* | 0.106 | (0.049, 0.164)* |
| Montana | -0.222 | (-0.299, -0.145)* | 0.061 | (0.017, 0.105)* | 0.029 | (-0.021, 0.079) |
| Wyoming | 0.212 | (0.054, 0.369)* | 0.271 | (0.182, 0.360)* | 0.153 | (-0.007, 0.313) |

\*p<0.05 †Note: Adjusted for below poverty variable, the percentage of people without high school diploma, the percentage of minority, the percentage of people with a disability, the percentage of crowding, the percentage of people 65 years and older, and for weeks.

## DISCUSSION

From March 2020 through January 2021, North Dakota, South Dakota, Idaho, Montana, and Wyoming have taken various approaches on implementing both policy and health guidelines to reduce the community transmission of SARS-CoV-2. Utilizing both the 7-day-sliding window *R*_*t*_ and policy change *R*_*t*_, we estimate the change in transmission potential over time and after each major policy change. Throughout the initial spring and summer surges across the U.S. coastal and major metropolitan areas, limited community spread of the virus was observed in the five states. Although we did not observe a corresponding surge in reported case numbers in South Dakota in August, the 7-day-sliding-window *Rt* has a small peak, suggesting there was community transmission related to Sturgis Motorcycle Rally, that was held in Meade County, South Dakota, August 7-16, 2020. Dave et al.,^19^ and Carter et al.,^20^ assessed the nationwide transmission of SARS-CoV-2 following the Sturgis Motorcycle Rally and identified it as a superspreading event. However, entering fall 2020, this region began to see an uptick in case counts ultimately leading a peak seen in late November. During the summer, reopening orders were given and between mid-August and September 1^st^, all five states resumed face-to-face K-12 schooling. Many of the universities also began in-person instruction. By September 7^th^ the *R*_*t*_ in all five states had increased by an average of 11.55%.

This paper used 7-day sliding window *R*_*t*_ estimates and policy change *R*_*t*_ estimates to assess and evaluate the various non-pharmaceutical interventions and policy changes at the state level for North Dakota, South Dakota, Idaho, Montana, and Wyoming. A few studies have accessed the effectiveness of mitigation measures, especially regarding face mask mandate and school reopening.^21-23^ Among the five states, North Dakota, Montana and Wyoming implemented the mask mandate and followed with the *R*_*t*_ reduction, which echoed with the current existing body of research on the efficacy of face masking in preventing the transmission of SARS-CoV-2.^24-26^ However, South Dakota and Idaho also experienced the *R*_*t*_ reduction without adopting the mask mandate, perhaps due to voluntary adoption of facemasks, which will require further investigation.

Furthermore, we observed an increase in *R*_*t*_ after schools (K-12 schools, colleges, and universities) reopened in South Dakota, Idaho, Montana, and Wyoming, except North Dakota. The detailed *R*_*t*_ description for all five states regarding school reopening are provided in **Appendix C**. School closure and suspended in-person instruction were considered to be one of the major public health interventions, and its effectiveness was widely discussed and researched. Our research results suggest that school reopening is correlated with an increase in *R*_*t*_ estimates; however, the results contradicted with the existing studies that the in-person instruction posed a low risk of transmission of SARS-CoV-2.^27-29^ This might reflect a concurrent change in social contact patterns when parents returned to workplaces after their children went back to schools. School closure as a mitigation strategy requires further research on its effectiveness since it is highly related to school-aged children, adolescents, and young adults’ physical health, mental health, and quality of life.^30^

North Dakota had a higher cumulative case count per capita in the more densely populated counties; however, the reason of why North Dakota was the only state of the 5 states under study that had statistically significant slopes (*m*) at all four assessed time points requires further investigation. Additional analysis found that 8 out of 20 slopes were very close to 0 (0<*m*<0.1), and the per capita cumulative case count did not vary by different population sizes.

When adjusting for SVI variables, our study found a positive significant association between log_10_-transformed per capita weekly incident case count and log_10_-transformed population size for Idaho, North Dakota, and South Dakota, with borderline significance for Wyoming, and no significance for Montana. Concretely, a 10-fold increase in population size was associated with a 0.106 increase in log_10_-transformed per capita incident case count in Idaho, 0.195 increase in North Dakota, and 0.127 increase in South Dakota. This positive significant association means that high-population counties have a higher per capita incident case count than low-population counties. This finding is supported by Wong & Li^31^ study where U.S. counties with more people were more likely to have larger numbers of cases especially in late Spring and early Summer 2020, or by McLaughlin et al.^32^ who emphasized the association of COVID-19 case rate with densely populated counties, urban counties and crowded housing.

## LIMITATIONS

There are some limitations in our study. First, upticks in cases may be due to external events attracting large crowds, such as a July 4, 2020 outdoor rally by the then President Trump at Mt. Rushmore, South Dakota, and a motorcycle rally in Sturgis, South Dakota in August 2020. It is impossible to distinguish local cases that were associated with the Sturgis Motorcycle Rally from those that were not using the aggregate data. Others have found evidence suggesting that the motorcycle rally might be a superspreading event, leading to at least 649 cases nationally that were associated with transmission chains traced back to the event, and Meade County (where the Rally was) experiencing a faster rate of growth in case rate than the rest of South Dakota, a week after the close of the Rally.^19,20^ The latter event occurred just before school opening and its independent effect on *R*_*t*_ increase may be difficult to tease out. Second, the date of symptom onset or the date of (unobserved) infection was not available for our dataset. Only the date of report was available. To correct for the time lag, we shifted the epidemic curve by nine days in order to correct for the incubation period and the delay to test results. Third, our dataset lacked information to distinguish between local and foreign imported cases. However, such distinction was mostly important in the early stages of the epidemic and since April 2020 community transmission was the main driver for the epidemic. Therefore, we argue that the absence of this distinction would not have significantly affected *R*_*t*_ estimates since April 2020. Fourth, we used aggregated numbers of reported cases by political jurisdictions instead of separate data on different facilities or settings, while each setting might demonstrate a different dynamic than that of community transmission. Fifth, underreporting due to the limits on testing capacity was especially acute at the beginning of the pandemic. The majority of the states issued orders to increase testing capacities between March and April 2020, and thus overcome this challenge. However, mild and asymptomatic cases are unlikely to get tested and confirmed. Thus, state-reported data cannot be used to measure the extent to which asymptomatic spread has progressed during the pandemic. Lastly, government orders to undertake a mitigation practice, and actual compliance, may differ, which is a recognized limitation in the data.^33,34^

## CONCLUSIONS

In North Dakota, South Dakota, Idaho, Montana, and Wyoming, new cases of COVID-19 started to rise in November and peaked in November-December 2020. From March 2020 to January 2021, the *R*_*t*_ for North Dakota, South Dakota, Idaho, Montana, and Wyoming fluctuated around one (with a range of 0.5 to 1.5 starting from June). Various social distancing policies including stay-at-home order and closing businesses and other protective interventions such as mask requirements appeared to be associated with a reduction in the spread of SARS-CoV-2 and keeping the *R*_*t*_ at a low level in the states studied in this paper. On May 13, 2021, the Centers for Disease Control and Prevention updated their guidelines regarding face coverings indicating that fully vaccinated people do not have to wear masks or practice physical distancing except where required by federal, state, local, tribal, or territorial laws, rules, and regulations.^35^ Since individuals fully vaccinated with COVID-19 vaccine still have a small chance becoming infected, this updated guideline is controversial among some healthcare professionals and requires further investigation.^36^ While we await the vaccination of more people so that the herd immunity threshold can be reached, government agencies may consider the continuation of the above-mentioned policies to manage transmission and keep the *R*_*t*_ below 1.0.

## Data Availability

All the data used in this paper are publicly available, aggregated, data. The cumulative confirmed case count data are available at the New York Times GitHub data repository. The 2019 county-level estimated population data are available from the US Census Bureau.

## Abbreviation list

COVID-19: Coronavirus disease 2019
SARS-CoV-2: Severe acute respiratory syndrome coronavirus 2
SVI: Social Vulnerability Index
U.S.: United States

## Supplementary material

### Appendix A. Data cleaning: handling negative incident case counts

Dates with negative case counts were identified (i.e., when public health agencies made corrections to their cumulative case counts at a specific date or dates). Daily case counts were adjusted to remove negative counts that were created when states adjusted their positive cases; for instance, a state would record negative cases in order to account for a previous over-count. To remove these negative counts all data was reviewed, and negative counts were transformed into a zero count by removing this number from the previous days with positive cases until there was no discrepancy. We adjusted the negative daily case counts at both state and county levels in all five states in this study.

### Appendix B. Assessing power-law relationship using linear regression of log-transformed per capita cumulative case count and log-transformed population size

If we assume that a power-law relationship exists between the cumulative case count (C) and the population size (N), with an exponent g, i.e., C = N^g, and that per capita cumulative case count, A = C/N, then,

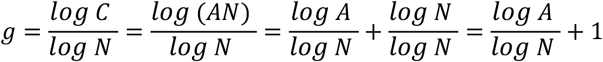

Thus,

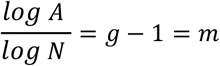

Where *m* is the slope of the regression line between the log_10_-transformed per capita cumulative case count and the log_10_-transformed population size.

### Appendix C. The detailed description for *R*_*t*_ in North Dakota, South Dakota, Idaho, Montana, and Wyoming

#### North Dakota

The daily reported cases of COVID-19 in North Dakota started to increase since July 2020, reaching a peak of 2,270 new cases in early November and 2,340 new cases in early December (Figure 3). The 7-day-sliding-window *R*_*t*_ stayed above 1 almost all year around and went below 1 in late May and in December. The governor’s executive orders and State Health Officers’ orders (Supplemental Table 2) throughout 2020 were chosen to create the policy change *R*_*t*_ throughout the year (Figure 3). From March 16, 2020 till April 4, 2020, the 7-day-sliding-window *R*_*t*_ decreased toward 1 but was still above 1 meaning that the promotion of physical distance was working. On April 18, 2020, the State Health Officer released the quarantine order for the Grand Forks county, and subsequently on May 14, 2020, they issued the testing order for Grand Forks county. In late May, the 7-day-sliding-window *R*_*t*_ fell below 1 where the coverage of the testing order expanded further. From June till November both the 7-day-sliding-window *R*_*t*_ and policy change *R*_*t*_ maintained a level above 1 which corresponded to the lifted quarantine order, the rescinded close contact quarantine order, and the permission of health care workers who are asymptomatically infected with SARS-CoV-2 to work in care facilities. From mid-November, the 7-day-sliding-window *R*_*t*_ and policy change *R*_*t*_ fell below 1, which corresponded to the release of the first mask order, and the allowance of team practice, bars and restaurant to resume normal hours.

#### South Dakota

From March 1, 2020 through January 1, 2021 South Dakota had 103,318 confirmed COVID-19 cases cumulatively. The daily reported cases of South Dakota had one late surge, peaking in November 2020 (Figure 3). Within South Dakota, there were eight policies or recommendations for COVID-19 precautions were implemented, five major ones can be found in Supplemental Table 2. The 7-day-sliding window *R*_*t*_ estimate fell below 1 on April 4, 2020 following the closure of the Smithfield pork processing plant in Sioux Falls, Minnehaha, South Dakota.^1^ The policy change *R*_*t*_ estimate remained below 1 until April 28, 2020 when South Dakota implemented their *Back to Normal Plan* which largely focuses on good hygiene practices and limiting persons in large numbers. From August 13 till September 25, 2020 the policy change *R*_*t*_ estimates increased to above 1 when South Dakota K-12 school reopened in person. The 95% credible intervals (CrI) of 7-day-sliding window *R*_*t*_ estimates in March 2020 were wide due to the small numbers of daily new cases. Meanwhile, during the periods between April 28, 2020 and August 13, 2020 and between September 25, 2020 and January 10, 2021 there were very few policy changes; the policy change *R*_*t*_ estimates may not accurately depict the true *R*_*t*_ estimates during those periods.

#### Idaho

From March 1, 2020 to January 1, 2021, Idaho’s daily reported case number revealed two major surges of new cases in July and November 2020 (Figure 3). The policy change *R*_*t*_ estimates based on statewide policy changes demonstrate the effectiveness of the non-pharmaceutical interventions (face masking, social distancing, prohibited gathering of more than 10 people, etc.) of controlling the spread of COVID-19 within the state of Idaho. The policy change *R*_*t*_ estimates dropped below 1 after the Statewide Stay Home order was in place between March 25, 2020 and April 30, 2020. The statewide policy change *R*_*t*_ estimates increased above 1 between May 1, 2020 and October 26, 2020, with the Stay Healthy Order relaxation of the controlling measures from stage 1 (encourage vulnerable individuals to stay at home, avoid public and private gathering, and avoid and minimize non-essential travels) to stage 4 (open all business, allow public gathering with any size and resume non-essential travel). With the Stay Healthy Order stage 3 in effect on October 27, 2020, the policy change *R*_*t*_ estimates dropped again and continued to drop below one, after the Stay Healthy Order moved from stage 3 back to stage 2 with more restrictions on November 14, 2020. Overall, Idaho demonstrated an extensive community transmission of SARS-CoV-2 transmission in the study period.

#### Montana

The number of incident cases in Montana had a surge in November 2020 with up to 1,622 daily cases in mid-November (Figure 3). The 7-day-sliding-window *R*_*t*_ curve and the policy change *R*_*t*_ curve show some significant changes over time parallel with implementing different COVID-19 related policies (Figure 3). There was a drastic drop in the 7-day-sliding-window *R*_*t*_ estimates after closing businesses and the *stay-at-home* orders at the end of March, leading to *R*_*t*_ estimates under 1 from late March until early May. An increase in 7-day-sliding-window *R*_*t*_ was observed in the first days of May as the state lifted the *stay-at-home* order, reopened schools and started the first phase of reopening businesses (such as some retails, restaurants, casinos, and bars). There was a decrease in policy change *R*_*t*_ estimates in mid-July after the statewide mask requirement was applied, with a slight rise in September, probably due to the start of schools’ Fall semester. In November after applying new restrictions on gatherings and also placing time and capacity limitations for bars and restaurants, the policy change *R*_*t*_ estimates dropped again below 1 and remained low, mostly till late December.

#### Wyoming

From March 1, 2020 to January 1, 2021, Wyoming’s daily reported case number revealed one major surge of new cases in October and November 2020 (Figure 3). The policy change *R*_*t*_ estimates based on statewide policy changes demonstrated the necessity of the non-pharmaceutical interventions (face masking, social distancing, reduction in the size of public gathering, etc.) of controlling the spreading of COVID-19 within the state of Wyoming. For instance, the *R*_*t*_ estimates stayed around and above 1 while the state opened up all businesses without face mask mandate and public gathering restrictions. The 7-day-sliding-window *R*_*t*_ estimates then dropped below 1 after the Seventeenth Continuation and Modification Statewide Public Health Order regarding the public gathering with no more than 25 people on November 19, 2020. With the first public health order regarding mandatory face covering in effect on December 7, 2020, the policy change *R*_*t*_ estimates maintained below one. Overall, Wyoming demonstrated an extensive community transmission of SARS-CoV-2 transmission in the study period.

### Appendix D: Detailed *R*_*t*_ description for North Dakota, South Dakota, Idaho, Montana, and Wyoming regarding School reopening

#### North Dakota

North Dakota decided to reopen schools and universities on June 1st, 2020.^2^ There are around 21 universities in the state located in Cass, Grand Forks, Ward, Burleigh, Stark, Stutsman, Trail, Mountrail, Bottineau, Benson, Barnes, Pierce, Dickey, Sioux, and Rolette counties.^3^ During the year of 2020, in most of the counties, the case number starts to increase at the end of the summer, peaking in November. Concretely, in Ward, Burleigh, Stark, Trail, and Sioux counties, the reported cases started to increase in July; in Cass, Grand Forks, Stutsman, Barnes counties, the reported cases started to increase in August; in Mountrail, Bottineau, and Benson counties the reported cases started to increase in September, while for the Pierce, Dickey, and Rolette counties, the daily reported case count started to increase in October, peaking in November. For most of the counties, the *R*_*t*_ stays at one during all the summer-fall period, except for some fluctuations (above and below one) in Grand Forks, and an *R*_*t*_ greater than one for Burleigh (where the state capital is located), and Dickey or Pierce (over one till the end of October). This description may be related to college students returning to school because the increase in the daily reported case count corresponds to the start of the fall semester in North Dakota. However, after November, new cases infected with SARS-CoV-2 and *R*_*t*_ starts to fall, which correspond to the issued mask order.

#### South Dakota

Some South Dakota schools began to offer in-person classes in June, but most schools did not begin face-to-face instruction for the mid-August fall semester.^4^ The state is home to four technical institutes, six state universities, six tribal colleges and universities, and six private colleges and universities; of these, six located in Minnehaha county, three in Pennington county, two schools located in Davison and Brown counties respectively, and one in Lawrence, Lake, Codington, Yankton, Oglala Lakota, Todd, Roberts, Brookings, and Clay counties respectively.^5^ Following the mid-August openings nine of these counties had an *R*_*t*_ that remained above 1 for between 7-14 days and four had *R*_*t*_ values that fluctuated both below and above 1. However, it is important to note that many of these counties have a very small population with small daily incident case count, which resulted in wide CrIs for *R*_*t*_ estimates around this time period. This initial increases in *R*_*t*_ and case count might be related to the return of students to in-person learning. In most counties with colleges and universities the epidemiologic increases seen in mid-August transitioned into the overall state-level fall surge that peaked around November. It is also noteworthy that the Sturgis Motorcycle Rally was held in Meade County, South Dakota, August 7-16, 2020, which coincide with the school reopening on August 13, 2020. Therefore, it is difficult to attribute the increase in Rt to a single policy decision (schools reopening) or a single mass gathering (Sturgis Motorcycle Rally). Both could have been factors involved; please refer to the discussion in the Limitations section in the main text.

#### Idaho

Idaho schools opened on-site instruction for the fall 2020 semester with a framework on decision making based on four different levels of community transmission.^6^ The state is home for four public colleges and 4-year universities, four regional community colleges, and eight private colleges and universities. Of these, four located in Boise, two in Moscow, two in Nampa, two in Idaho Falls, and one in Rexburg, Caldwell, Meridian, Twin Falls, Pocatello, Lewiston, McCall, and Coeur d’Alene respectively.^7^ Following the mid-August openings, Boise County had an *R*_*t*_ fluctuated below and above 1 but mainly above 1; however, the daily case counts were very small, less than 15, therefore the CrI of *R*_*t*_ was wide. Boise County is a college town for four colleges and universities. These small daily case counts may relate to effective non-pharmaceutical interventions or issues of underreporting.

#### Montana

Montana was one of the 2 states that resumed in-person instructions in schools in the academic year 2019-2020, on May 7, 2020.^8^ As seen in Figure 3, *R*_*t*_ rose above one from early May and stayed high until mid-July which reflected the impact of this policy in the state, in contrast to most of the other states of the county. There are more than 20 colleges and universities in Montana, with approximately 40,000 students, in Gallatin county, Yellowstone county, Hill county, Missoula county, Silver Bow county, Beaverhead county, Dawson county, Flathead county, Custer county, Lewis and Clark county, Cascade county, and tribal colleges in the counties of Blaine, Glacier, Rosebud, Roosevelt, Big Horn, Lake and Hill.^9^ Fall semester 2020 started in the second half of August in almost all of these academic institutions with online and on campus courses, using new guidelines to improve safety of students. No significant change in *R*_*t*_ was observed in these counties after the Fall openings.

#### Wyoming

Wyoming K-12 schools, colleges and universities, and trade schools were reopened in limited on-site instructions on May 7, 2020, and later fully opened for face-to-face on-site instruction on June 10, 2020.^10^ The state is home for one 4-year university, eight public colleges, one tribal college, and two private colleges. Except for the University of Wyoming and Wyoming Technical Institute located in Laramie, the rest of the colleges scatter across different counties.^11^ Following the reopening in June, Laramie County had an *R*_*t*_ spike of 4 and *R*_*t*_ fluctuated below and above 1 but mainly above 1 between June and December, and *R*_*t*_ dropped below 1 between December and January. The largest daily case count was 246 on November 14, 2020. The trend of *R*_*t*_ and case count increase might be related to the school resumed face-to-face instruction in June and following fall semester.

### Appendix E

Linear regression between log_10_-transformed per capita cumulative case number (ccn) and log_10_-transformed population size by county for North Dakota, South Dakota, Idaho, Montana, and Wyoming, on June 30^th^, August 31^st^, October 31^st^, and December 31^st^, 2020 (date of report) are presented in Figures S1-S5.

**Figure S1.**
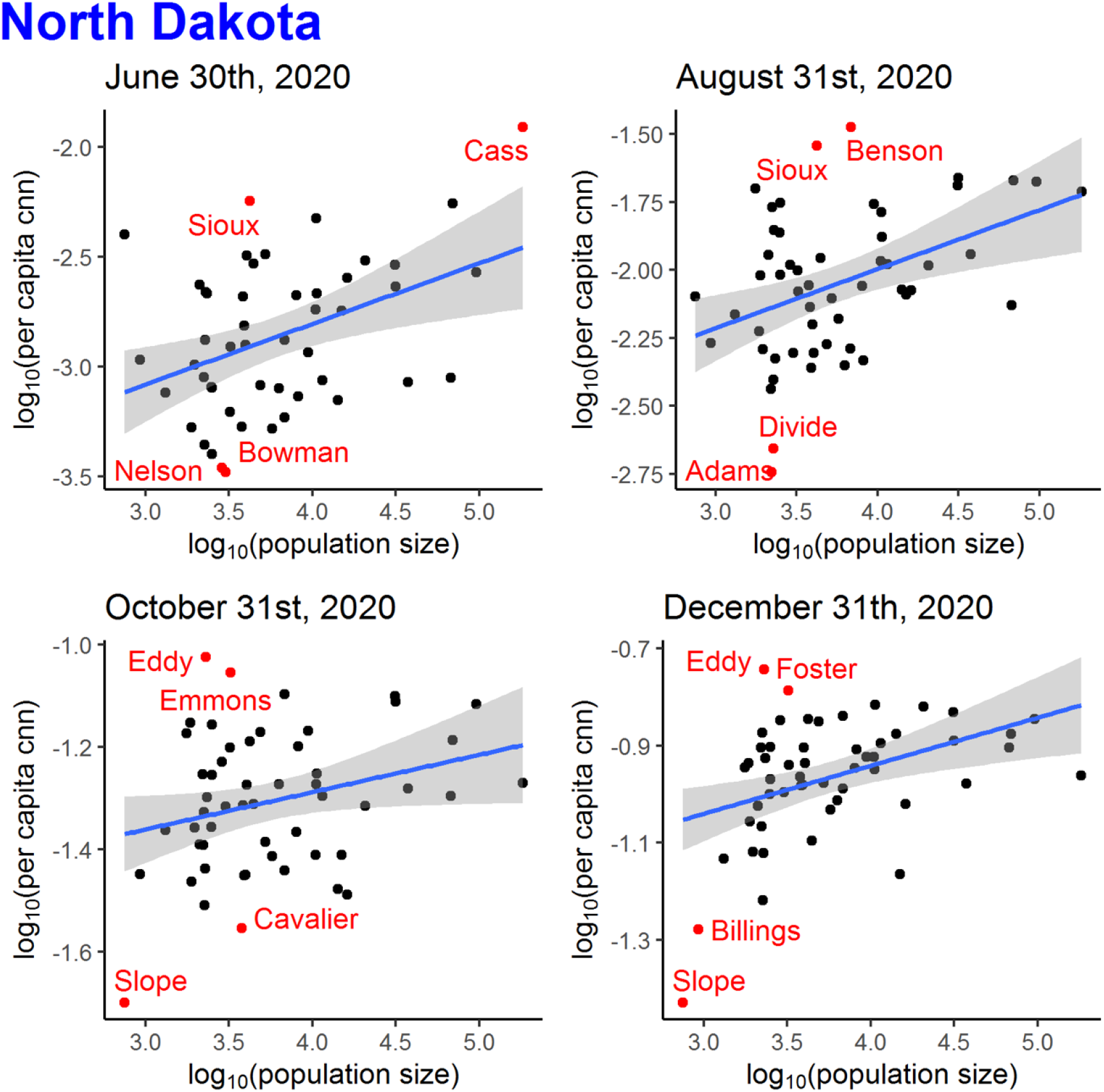
Linear regression between log_10_-transformed per capita cumulative case number (ccn) and log_10_-transformed population size by county for North Dakota (53 counties), on June 30^th^, August 31^st^, October 31^st^, and December 31^st^, 2020 (date of report). Counties that were outliers (defined as below 2.5 percentile or above 97.5 percentile of the distribution of the log_10_-transformed per capita ccn) were marked in red.

**Figure S2.**
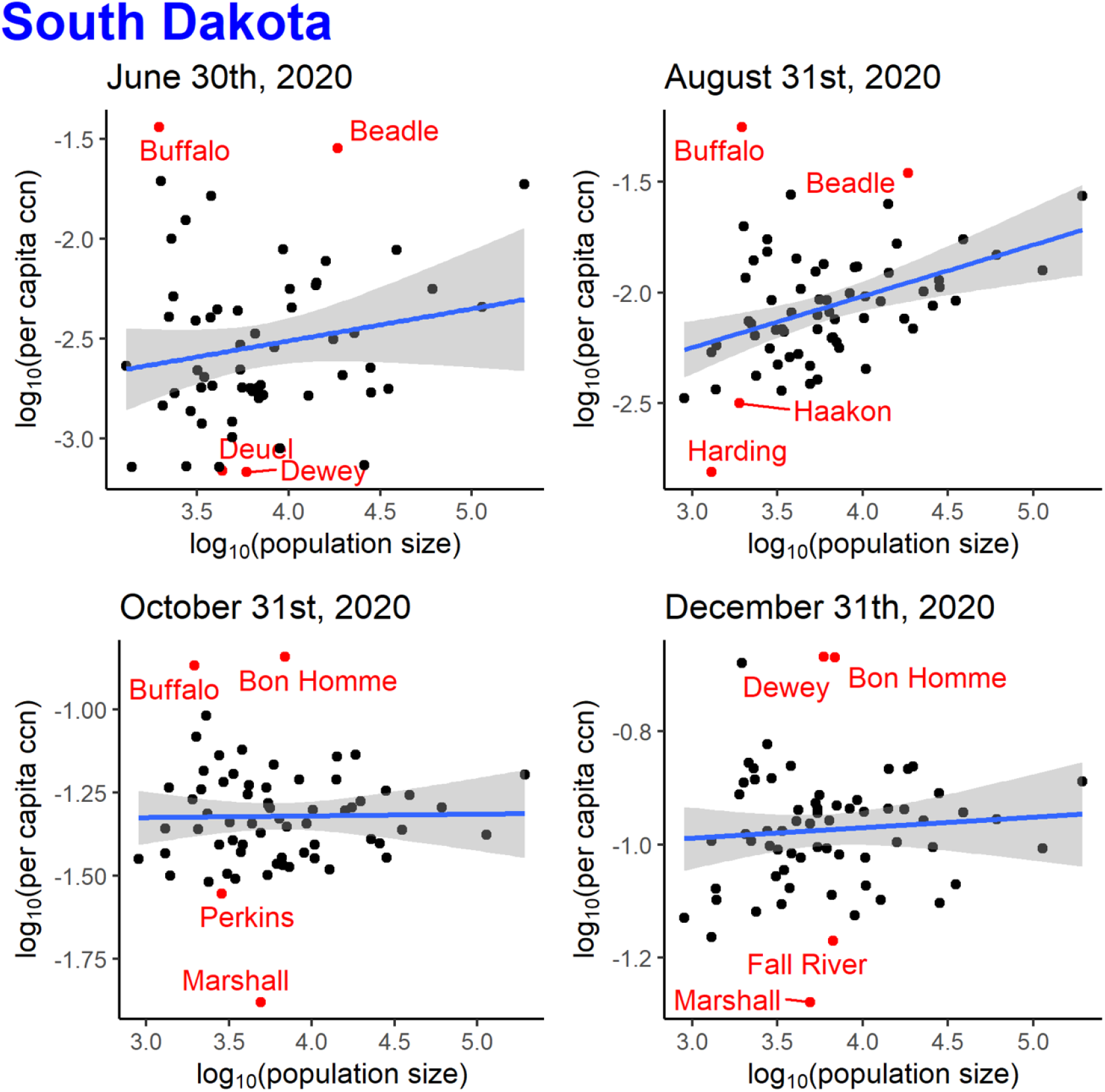
Linear regression between log_10_-transformed per capita cumulative case number (ccn) and log_10_-transformed population size by county for South Dakota (66 counties), on June 30^th^, August 31^st^, October 31^st^, and December 31^st^, 2020 (date of report). Counties that were outliers (defined as below 2.5 percentile or above 97.5 percentile of the distribution of the log_10_-transformed per capita ccn) were marked in red.

**Figure S3.**
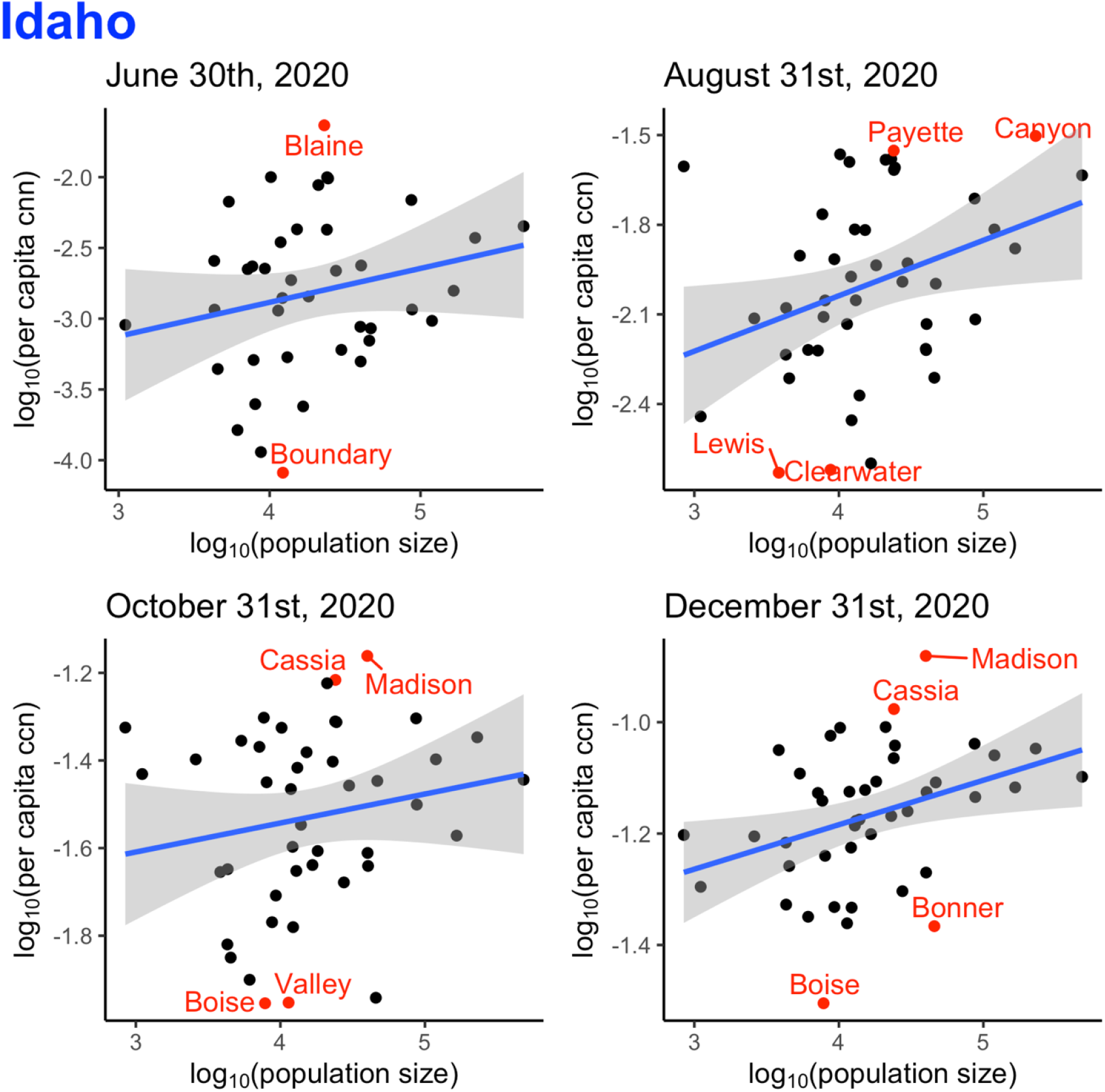
Linear regression between log_10_-transformed per capita cumulative case number (ccn) and log_10_-transformed population size by county for Idaho (44 counties), on June 30^th^, August 31^st^, October 31^st^, and December 31^st^, 2020 (date of report). Counties that were outliers (defined as below 2.5 percentile or above 97.5 percentile of the distribution of the log_10_-transformed per capita ccn) were marked in red.

**Figure S4.**
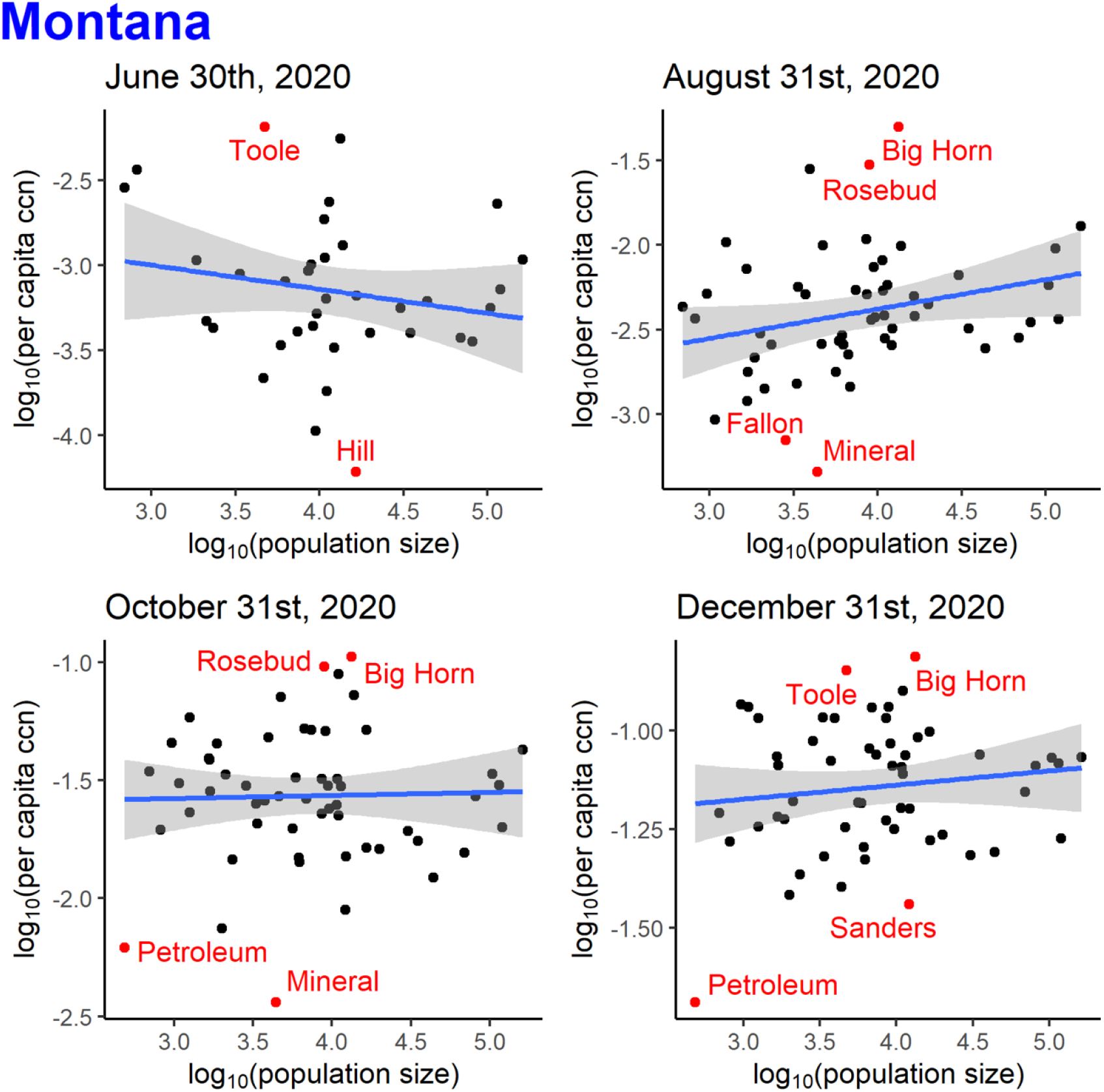
Linear regression between log_10_-transformed per capita cumulative case number (ccn) and log_10_-transformed population size by county for Montana (56 counties), on June 30^th^, August 31^st^, October 31^st^, and December 31^st^, 2020 (date of report). Counties that were outliers (defined as below 2.5 percentile or above 97.5 percentile of the distribution of the log_10_-transformed per capita ccn) were marked in red.

**Figure S5.**
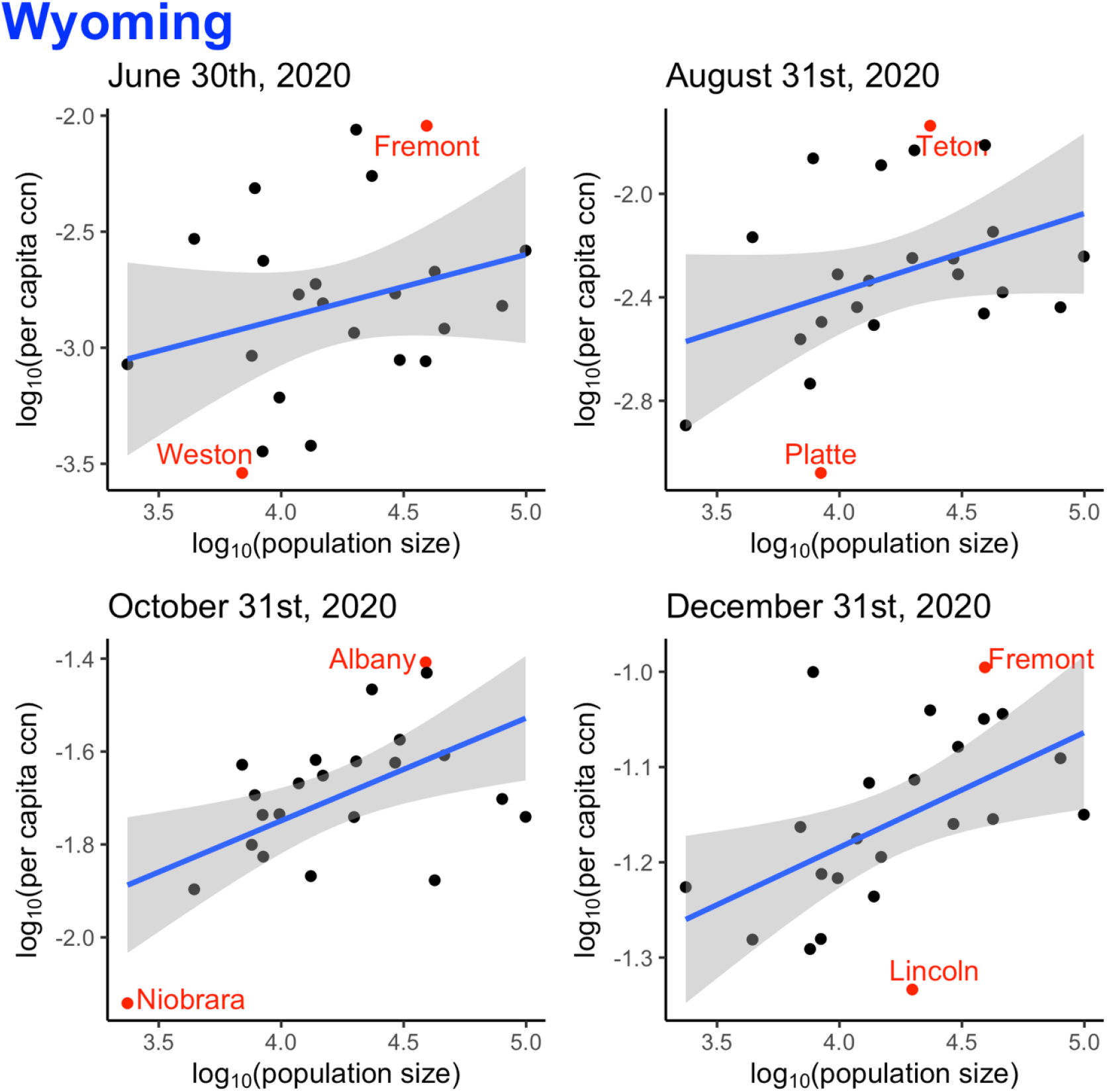
Linear regression between log_10_-transformed per capita cumulative case number (ccn) and log_10_-transformed population size by county for Wyoming (23 counties), on June 30^th^, August 31^st^, October 31^st^, and December 31^st^, 2020 (date of report). Counties that were outliers (defined as below 2.5 percentile or above 97.5 percentile of the distribution of the log_10_-transformed per capita ccn) were marked in red.

**Supplemental Table 1.**
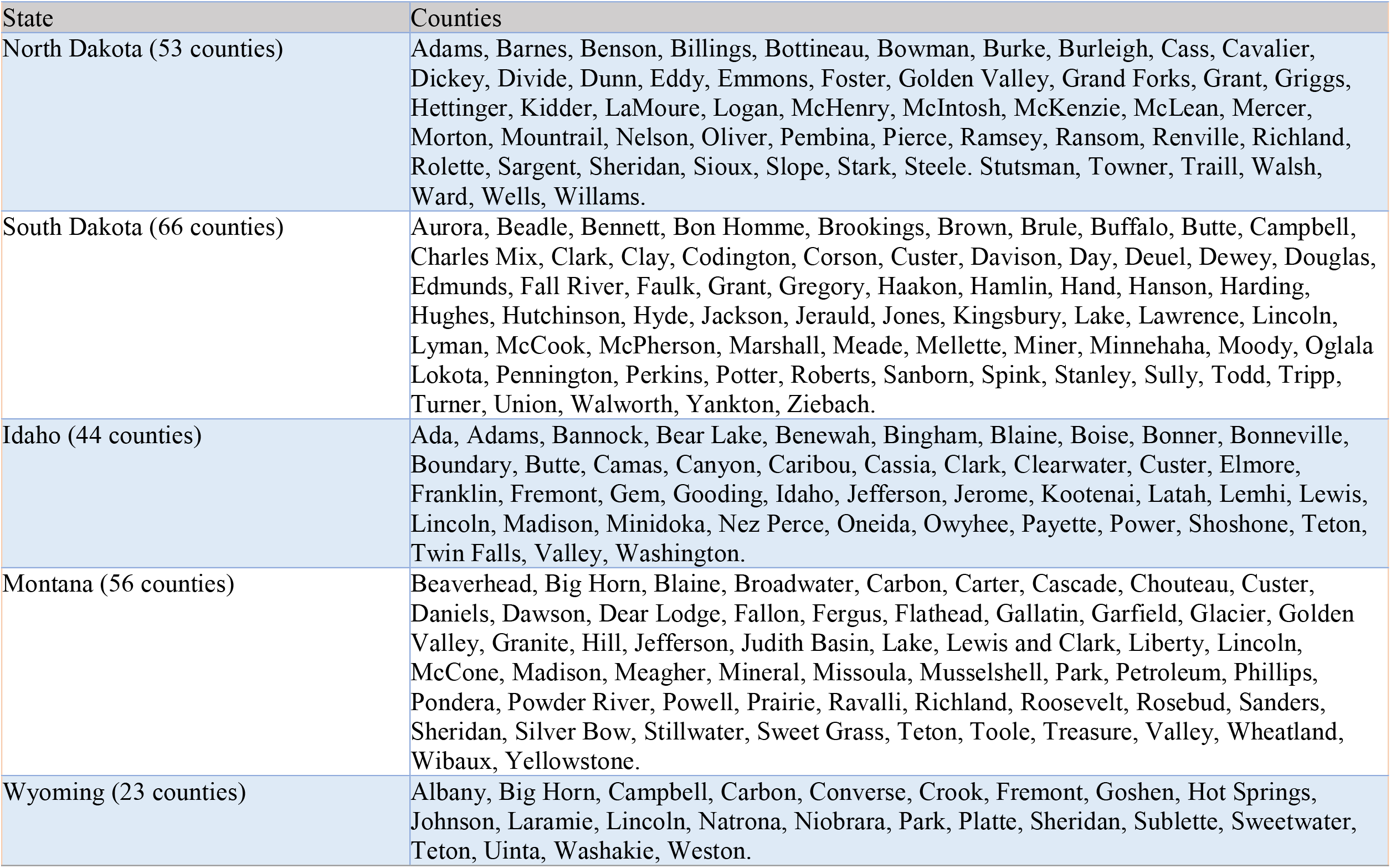
List of counties in North Dakota, South Dakota, Idaho, Montana, and Wyoming (organized by state).^12^

**Supplemental Table 2:** Control measures implemented by state government agencies in North Dakota, South Dakota, Idaho, Montana, and Wyoming, USA

|  | Label | Date<br>(mm/dd/yyyy) | Implemented measure(s) |
| --- | --- | --- | --- |
| North Dakota | A | 03/16/2020 | Executive Order 2020-04.1. The governor amended school closure order for certain schools and programs.<sup>13</sup> |
|  | B | 03/30/2020 | Executive Order 2020-10.1. The governor enabled remote participation for public meetings to promote physical distancing. He amended school facilities closure to accommodate child care needs and suspended deadlines for livestock auctions, pesticide applicators, and public libraries.<sup>14</sup> |
|  | C | 05/26/2020 | Testing Order \| Long Term Care. The State Health Officer Order for testing for 2019-nCoV/COVID-19 as a Disease Control Measures to prevent the spread 2019-nCoV/COVID-19.<sup>15</sup> |
|  | D | 09/21/2020 | Order #2020-02.4 \| Lifted Travel Quarantine Order. The State Health Officer Order lifted the 14-day quarantine order for those returning from international travel.<sup>16</sup> |
|  | E | 11/13/2020 | Order #2020-08 \| Mask Order. The mask order required face coverings in indoor businesses and indoor public settings, as well as in outdoor business and public settings when it was not possible to maintain physical distancing.<sup>17</sup><br>Executive Order 2020-43. The governor enacted mitigation measures for food service, events, sports, and extracurricular activities.<sup>18</sup> |
|  | F | 12/21/2020 | The governor amended the order to allow restaurants and bars to resume normal hours.<sup>19</sup> |
| South Dakota | A | 3/13/2020 | State of emergency declared, executive state government workers given work from home order and restricted out of state travel (EO# 2020-05); K-12 schools closed for the rest of the year; higher education encouraged to close campus; and long-term care facilities implemented visitor restrictions.<sup>20</sup> |
|  | B | 4/6/2020 | All residents shall review and practice CDC COVID-19 guidelines; all employers, local, and municipal government shall implement CDC COVID-19 guidelines to encourage telework, social distancing, and reduced numbers; all healthcare workers shall prepare for increased COVID-19 cases (EO# 2020-12)<sup>21</sup>; stay-at-home order except for essential workers and essential outings for all Minnehaha and Lincoln “vulnerable individuals” (EO# 2020-13).<sup>22</sup> |
|  | C | 4/28/2020 | All citizens shall implement “back to normal plan” leaving COVID-19 precautions up to each resident, employers, and schools; ended Minnehaha and Lincoln “vulnerable individuals” stay-at-home order; (EO# 2020-20).<sup>23</sup> |
|  | D | 8/13/2020 | All K-12 schools were mandated by the South Dakota Governor to start in-person classes again.<sup>24</sup> |
|  | E | 9/25/2020 | Long-term care facilities began to relax visitor restrictions.<sup>24</sup> |
| Idaho | A | 3/25/2020 | Statewide stay-home order. This isolation order required Idaho residents to stay and work from home as much as possible while ensuring all essential services and business remain available. This isolation order did not prohibit outdoor activity such as walking, hiking, running, and biking, but a safe distance of six (6) feet must be kept between those who do not live in the same household.<sup>25</sup> |
|  | B | 5/1/2020 | Stay Healthy Order-Stage 1: Businesses and governmental agencies resumed operations at physical locations in the state of Idaho except for those businesses identified in this order. All businesses must adhere to the social distancing and sanitation requirement. Certain individuals entering Idaho required self-quarantine for 14 days. Gathering, both public and private, should be avoided. Non-essential travel should be avoided or minimized.<sup>26</sup> |
|  | C | 6/13/2020 | Stay Healthy Guidelines-Stage 4: Gathering of any sizes were allowed but should adhere to the physical distancing and sanitation requirements. Non-essential travel could resume.<sup>27</sup> |
|  | D | 10/27/2020 | Stay Health Order-Stage 3: Indoor and outdoor gathering, both public and private must adhere to the requirement identified in this order. Patrons of bars, nightclubs, and restaurants must remain seated. Face coverings were required at long-term care facilities.<sup>28</sup> |
|  | E | 11/14/2020 | Stay Health Order-Stage 2: Gathering of more than 10 people, both public and private, were prohibited, including attendance at extracurricular activities such as sporting events. People at increased risk for severe illness living in the state of Idaho were strongly encouraged to stay home and limit their movement outside of their place of residence.<sup>29</sup> |
| **Montana**<sup>30</sup> | A | 3/13/2020 | State of Emergency declared and directive implementing Executive Orders 2-2020 and 3-2020 and providing for measures to combat the spread of COVID-19 Novel Coronavirus. Court closure. |
|  | B | 5/7/2020 | School reopened. |
|  | C | 7/15/2020 | Statewide mask requirement required individuals to wear masks inside certain businesses and at outdoor gatherings of greater than 50 people where social distancing was not possible. |
|  | D | 9/1/2020 | Schools' Fall semester began. |
|  | E | 11/18/2020 | Additional mitigation measures. Gatherings were limited to 25 people when social distancing is not possible. Bars, restaurants, and casinos will have a 10 p.m. curfew every night and were limited to 50% capacity. |
| **Wyoming** | A | 3/13/2020 | Statewide Public Health Order #1. Closed all restaurants, bars, theaters, gymnasiums, childcare facilities, K-12 schools, colleges, universities, and trade schools, in the State of Wyoming, with certain exceptions. This Order was effective immediately and shall remain in effect until April 3, 2020.<sup>31</sup> |
|  | B | 4/28/2020 | Statewide Public Health Order, 3rd Continuation and Modification. This order authorized gymnasiums and childcare facilities to reopen under certain conditions. This order was effective on May 1, 2020 and shall remain in effect through May 15, 2020. Forbade gathering for 10 people or more.<sup>32,33</sup> |
|  | C | 6/10/2020 | Statewide Public Health Order, 6th Continuation and Modification. This order authorized K-12 schools, colleges, universities, and trade schools to provide on-site instruction to students and allow others to use of their facilities under certain conditions. Forbade gathering for 50 people or more.<sup>10,34</sup> |
|  | D | 7/13/2020 | Statewide Public Health Order, 8th Continuation and Modification. This order removed some conditions and restrictions that were applicable to restaurants.<sup>35,36</sup> |
|  | E | 10/14/2020 | Statewide Public Health Order, 13th Continuation and Modification. This order required patrons at different tables (at restaurants/bars) to be seated at least 6 feet apart and did not apply to booths, and patrons could be in groups up to 8 instead of 6. This order also authorized groups of attendees seated or standing together during an event in groups of 8 instead of 6. Forbade gathering for 50 people or more.<sup>37,38</sup> |
|  | F | 12/7/2020 | Statewide Public Health Order #4. Required face coverings in certain places, with exceptions. This order was effective on December 9, 2020, and shall remain in effect through January 8, 2021.<sup>39</sup> |

**Supplemental Table 3.**
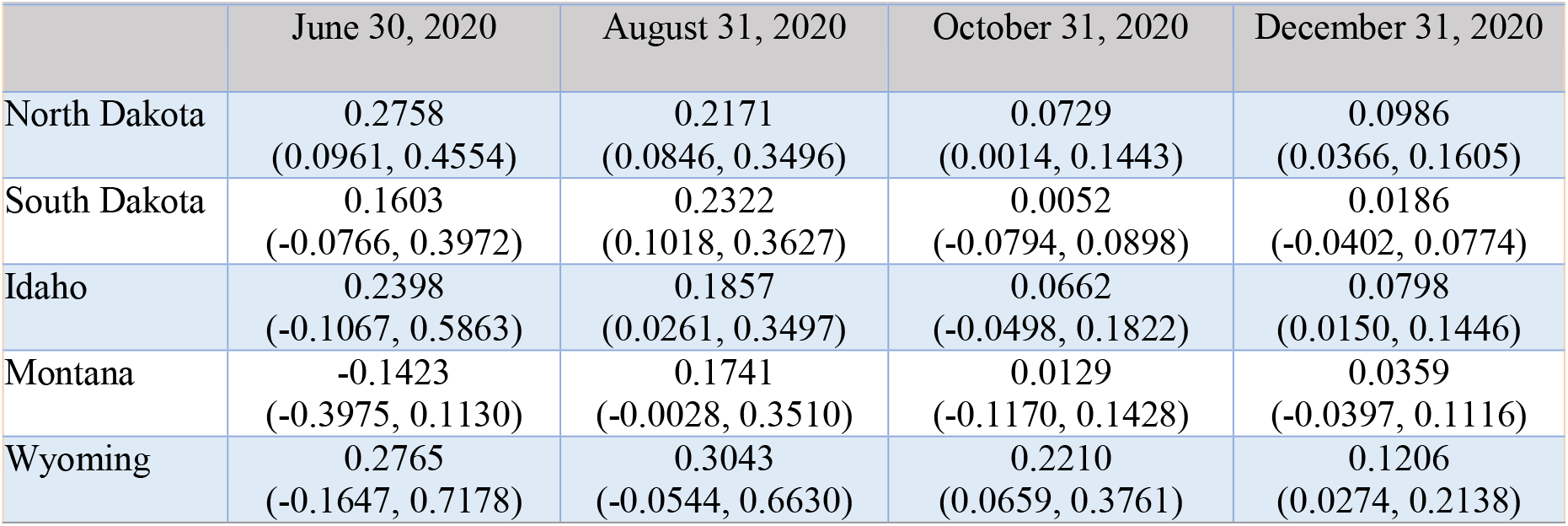
The slope (and 95% Confidence Intervals) of the regression line between log_10_-transformed per capita cumulative case count and log_10_-transformed population size, by states, North Dakota, South Dakota, Idaho, Montana, and Wyoming, USA, on June 30, August 31, October 31, and December 31, 2020.

|  | June 30, 2020 | August 31, 2020 | October 31, 2020 | December 31, 2020 |
| --- | --- | --- | --- | --- |
| North Dakota | 0.2758<br>(0.0961, 0.4554) | 0.2171<br>(0.0846, 0.3496) | 0.0729<br>(0.0014, 0.1443) | 0.0986<br>(0.0366, 0.1605) |
| South Dakota | 0.1603<br>(-0.0766, 0.3972) | 0.2322<br>(0.1018, 0.3627) | 0.0052<br>(-0.0794, 0.0898) | 0.0186<br>(-0.0402, 0.0774) |
| Idaho | 0.2398<br>(-0.1067, 0.5863) | 0.1857<br>(0.0261, 0.3497) | 0.0662<br>(-0.0498, 0.1822) | 0.0798<br>(0.0150, 0.1446) |
| Montana | -0.1423<br>(-0.3975, 0.1130) | 0.1741<br>(-0.0028, 0.3510) | 0.0129<br>(-0.1170, 0.1428) | 0.0359<br>(-0.0397, 0.1116) |
| Wyoming | 0.2765<br>(-0.1647, 0.7178) | 0.3043<br>(-0.0544, 0.6630) | 0.2210<br>(0.0659, 0.3761) | 0.1206<br>(0.0274, 0.2138) |

**Supplemental Table 4.** The adjusted linear regression model between log_10_-transformed per capita incident case count and log_10_-transformed population size for Idaho, March 2020 -January 2021*

| Variables | Parameter Estimate | Standard Error | t-Statistic | P-value |
| --- | --- | --- | --- | --- |
| Population | 0.106 | 0.030 | 3.606 | 0.0003 |
| Below poverty estimate percentage | 0.007 | 0.003 | 2.252 | 0.0244 |
| No high school diploma percentage | 0.005 | 0.006 | 0.827 | 0.4086 |
| Age 65+ percentage | -0.014 | 0.005 | -2.780 | 0.0055 |
| Population with disability percentage | 0.009 | 0.005 | 1.873 | 0.0613 |
| Minority percentage | 0.010 | 0.003 | 3.039 | 0.0024 |
| Crowding percentage | -0.030 | 0.010 | -2.921 | 0.0035 |

\*Note: adjusted for weeks

**Supplemental Table 5.** The adjusted linear regression model between log_10_-transformed per capita incident case count and log_10_-transformed population size for Montana, March 2020 - January 2021*

| Variables | Parameter Estimate | Standard Error | t-Statistic | P-value |
| --- | --- | --- | --- | --- |
| Population | 0.0290 | 0.026 | 1.136 | 0.2561 |
| Below poverty estimate percentage | -0.017 | 0.004 | -4.401 | 0.0000 |
| No high school diploma percentage | 0.008 | 0.004 | 2.196 | 0.0282 |
| Age 65+ percentage | -0.001 | 0.004 | -0.304 | 0.7614 |
| Population with disability percentage | -0.007 | 0.005 | -1.373 | 0.1698 |
| Minority percentage | 0.008 | 0.002 | 4.703 | 0.0000 |
| Crowding percentage | 0.015 | 0.008 | 1.820 | 0.0689 |

\*Note: adjusted for weeks

**Supplemental Table 6.**
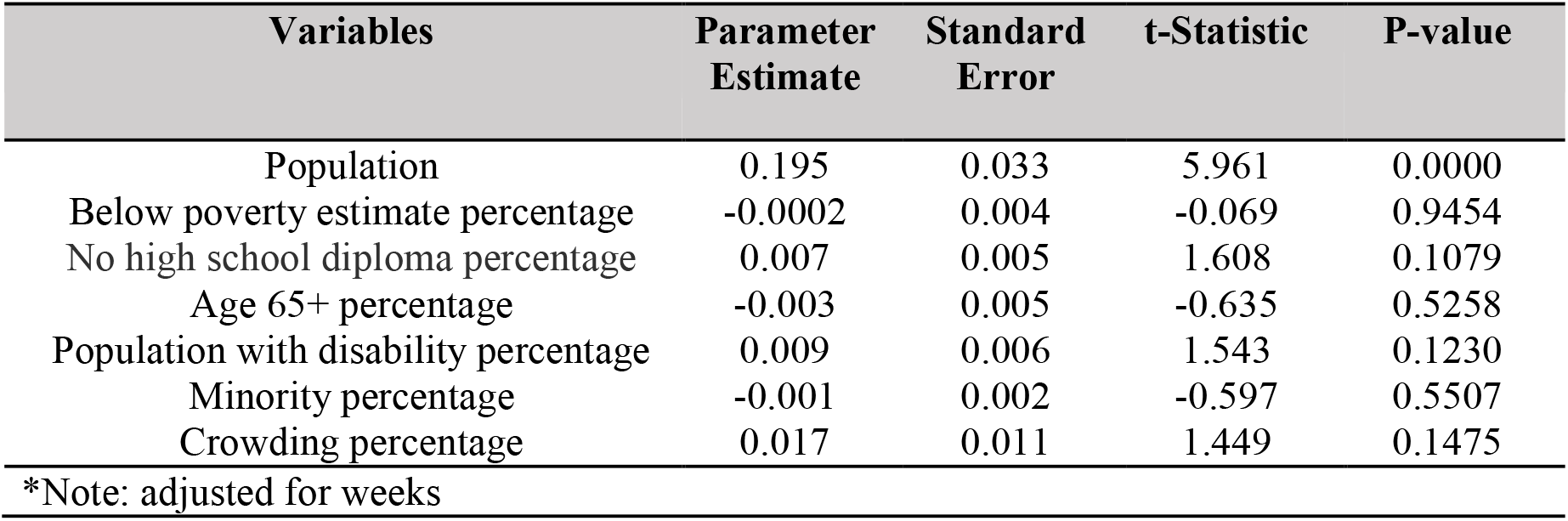
The adjusted linear regression model between log_10_-transformed per capita incident case count and log_10_-transformed population size for North Dakota, March 2020 - January 2021*

**Supplemental Table 7.**
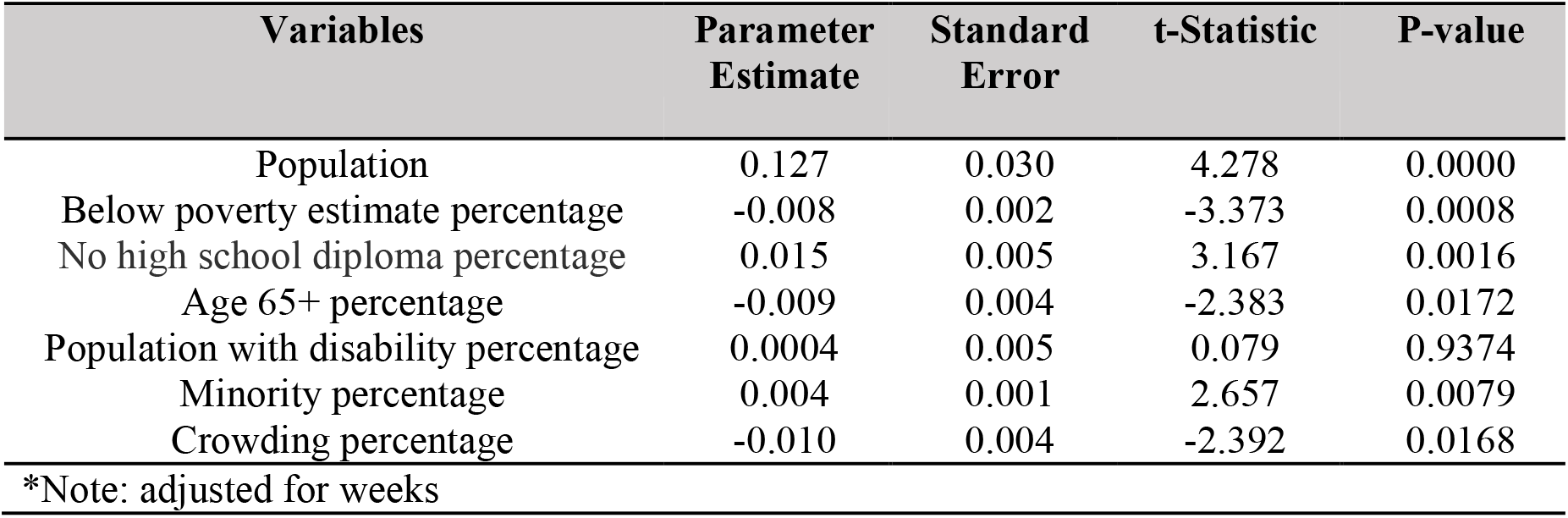
The adjusted linear regression model between log_10_-transformed per capita incident case count and log_10_-transformed population size for South Dakota, March 2020 - January 2021*

**Supplemental Table 8.** The adjusted linear regression model between log_10_-transformed per capita incident case count and log_10_-transformed population size for Wyoming, March 2020 - January 2021*

| Variables | Parameter Estimate | Standard Error | t-Statistic | P-value |
| --- | --- | --- | --- | --- |
| Population | 0.153 | 0.081 | 1.879 | 0.0606 |
| Below poverty estimate percentage | 0.0006 | 0.006 | 0.107 | 0.9145 |
| No high school diploma percentage | -0.026 | 0.012 | -2.206 | 0.0277 |
| Age 65+ percentage | 0.000 | 0.007 | 0.012 | 0.9901 |
| Population with disability percentage | 0.030 | 0.010 | 2.937 | 0.0034 |
| Minority percentage | 0.012 | 0.005 | 2.562 | 0.0106 |
| Crowding percentage | 0.090 | 0.018 | 5.017 | 0.0000 |

\*Note: adjusted for weeks

